# Influence of landscape patterns on the exposure of LASV across diverse regions within the Republic of Guinea

**DOI:** 10.1101/2022.02.28.22271612

**Authors:** Stephanie Longet, Cristina Leggio, Joseph Akoi Bore, Tom Tipton, Yper Hall, Fara Raymond Koundouno, Stephanie Key, Hilary Bower, Tapan Bhattacharyya, N’Faly Magassouba, Stephan Günther, Ana-Maria Henao-Restrapo, Jeremy S. Rossman, Mandy Kader Konde, Kimberly Fornace, Miles W. Carroll

**Author notes:** Corresponding author Address for correspondence: Prof Miles W. Carroll & Dr Stephanie Longet, Nuffield Department of Medicine, Wellcome Centre for Human Genetics, University of Oxford, Oxford, OX3 7BN, United Kingdom. Emails. Contributed equally to first authorship (Stephanie Longet, Cristina Leggio, Joseph Akoi Bore).

## Abstract

Lassa fever virus (LASV) is the causative agent of Lassa fever, a disease endemic in West Africa. Exploring the relationships between environmental factors and LASV transmission across ecologically diverse regions can provide crucial information for the design of appropriate interventions and disease monitoring. We measured LASV-specific IgG seropositivity in 1286 sera collected in Coastal and Forested Guinea. Our results showed that exposure to LASV was heterogenous between the sites. The LASV IgG seropositivity was 11.9% (95% CI 9.7-14.5) in Coastal site, while it was 59.6% (95% CI 55.5-63.5) in Forested region. Interestingly, exposure was significantly associated with age, with seropositivity increasing with age in the Coastal site. Finally, we also found significant associations between exposure risk to LASV and landscape fragmentation in Coastal and Forested regions. This study may help to define the regions with an increased exposure risk to LASV where a close surveillance of LASV circulation is needed.

## Introduction

Lassa virus (LASV) is an enveloped and negative-sense, single-stranded RNA virus belonging to the *Arenaviridae* family causing Lassa Fever (LF) in humans *(1)*. Initial clinical presentation is often non-specific and could be indicative of several other febrile illnesses *(2)*. The mortality rate is unclear due to inconsistent reporting practices and lack of diagnostic services in most endemic areas but estimated to 5-20% in hospitalised patients *(2) (3). Mastomys natalensis* (*M. natalensis*) is considered as the main natural reservoir, a commensal rodent and agricultural pest which aggregates in human dwellings and surrounding fields *(4)*. Recent studies have identified additional rodent reservoirs thus revealing the potential for a wider ecological niche for the virus *(5)*. Both zoonotic and non-zoonotic transmission mechanisms have been described *(6)*. Human LASV infections most commonly occur via infected rodent excreta, through contaminated food and inhalation of aerosols from rodent urine or droppings *(7) (8)*. Less commonly, the virus can be transmitted via nosocomial transmission if proper barrier nursing methods are not followed *(6) (9)*.

LASV is endemic in West Africa particularly in Nigeria, Sierra Leone, Liberia and Guinea *(10) (11)*. A published estimate on annual cases of LASV infections in West Africa range from 100,000 to 300,000, with 5,000 to10,000 estimated deaths per year and 80% asymptomatic infections. Further epidemiological studies are needed as these figures have been extrapolated from one longitudinal study conducted in Sierra Leone in 1987 *(12)*. Increasing evidence suggests LASV is widespread and substantially under-reported by passive surveillance systems *(13)*. Thus, characterising the distribution and transmission intensity of LASV in endemic areas is essential for the design of effective surveillance and disease control programmes.

It was observed that patients, including with mild symptoms, induced LASV-specific antibodies including long-lived IgG *(14)*. Consequently, serological methods can measure long-lived antibody responses and enable estimation of cumulative exposure to pathogens *(15)*. The analysis of age-stratified serological responses is a powerful tool to reconstruct historical patterns of transmission and distribution of previously unreported cases *(16) (17)*. Measurement of LASV-antibody responses has been used to detect circulation of LASV across Guinea. Lukashevich *et al* found 25-55% LASV NP-specific IgG positivity among inhabitants of tropical secondary forest and guinea savannah but only 4-7% positivity among inhabitants of mountainous and coastal areas *(18)*. Bausch *et al* investigated the epidemiology and clinical presentation of the Lassa fever in five Guinean hospitals in 2001. Lassa fever suspected cases were tested using an ELISA platform for antibody detection and the analysis confirmed Lassa fever in 22 (7%) of 311 suspected cases. An additional 43 people (14%) had LASV-specific IgG antibodies, indicating past exposure to LASV *(19)*. Another study aiming to estimate the prevalence of Lassa IgG in human population of rural area of Guinea including Guéckédou, Lola and Yomou (Southern region) indicated 12.9% and 10.0 % IgG positivity in rural and urban areas, respectively *(7)*. While these studies describe the historical circulation of LASV across Guinea, these studies were conducted in the 90’s and early 2000’s. The current burden and the distribution of LASV in Guinea is largely unknown. Furthermore, to our knowledge, there are no studies which have estimated transmission intensity from serological data in Guinea and analysed the environmental risk factors. This study aimed to characterise LASV exposure in populations in two ecologically diverse regions of Guinea: the coastal Basse-Guinee region and the forested interior regions of Macenta prefecture and Guéckédou. Here, we describe LASV-specific IgG antibody levels within these populations, estimate transmission intensity using age-stratified antibody responses and explore associations with demographic and environmental risk factors.

## Materials and Methods

### Ethical approval

Basse Guinea site (Coastal site): The study was approved by the London School of Hygiene and Tropical Medicine Ethics committee (Ethics Ref: 17429) and the Guinean Ministry of Health (MoH). Macenta site: Ethical approval for research projects in Macenta and Guéckédou (Forested site) on human volunteers was obtained from the ‘Comité national d’éthique pour la recherche en santé’ (CNERS) of Republic of Guinea on the 16^th^ of March 2017 (012/CNERS/17).

### Study sites

The samples used in this study were initially collected as part of three separate studies focusing on three distinct sites within two geographically diverse regions of Guinea. Basse-Guinee is the coastal region of Guinea, extending for 30-40 miles inland from the Atlantic Ocean to the coastal plains (**Figure 1A**). Macenta prefecture and the city of Guéckédou are located to the south-east of the country, close to the border with Liberia and Sierra Leone, within the forested region of Guinea (**Figure 1A**).

**Figure 1:**
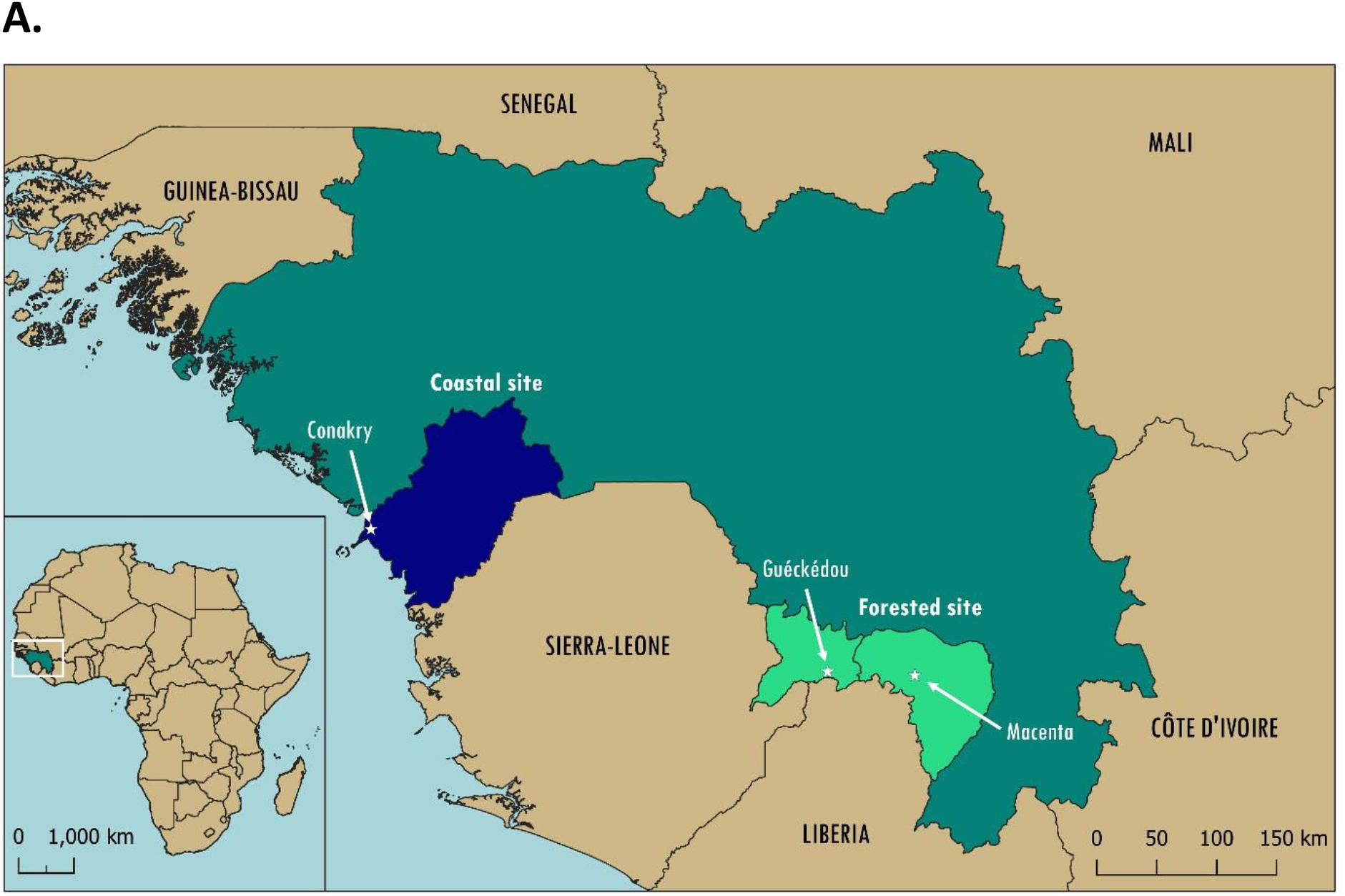
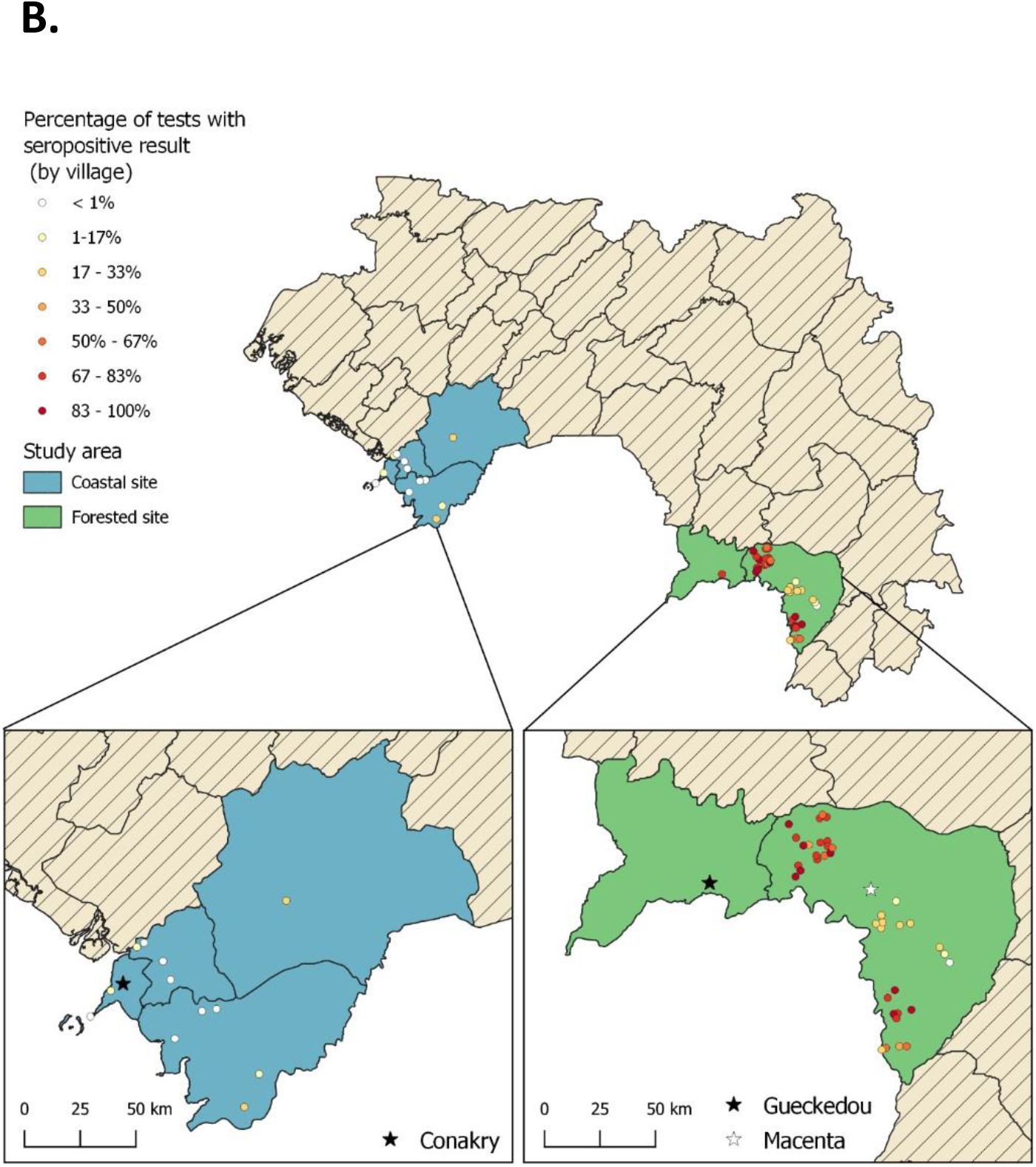
**A)** Map of West Africa and Guinea showing the study sites: Coastal and Forested sites. **B)** LASV-specific IgG seropositivity (%) by village across the Coastal and Forested sites.

### Study participants

Samples from Basse-Guinee were collected in 2016 as part of a clinical trial evaluating the safety and immunogenicity of the VSV-EBOV Ebola Vaccine in partnership with the Guinean MoH, the World Health Organisation (WHO) and Public Health England (PHE). The samples used for our study were taken prior to vaccination. For this study site, 702 samples were randomly selected from the PHE/WHO vaccine trial master database by using the Excel random function and tested for the presence of LASV-specific IgG. Information on volunteers age, sex and village of residence provided at the time of vaccination was used. The selected samples, (participants 6-99 years of age) originated from 7 prefectures within Basse-Guinee, encompassing 24 subprefectures and 29 villages/neighbourhoods.

Samples from Macenta region were collected from February to December 2017 as part of a sero-epidemiology study on Ebola virus which included both affected and non-affected villages during the 2014-2016 Ebola Virus Diseases (EVD) outbreak *(20)*. In total, 517 participants (17-90 years of age) living in 44 villages located in 7 subprefectures were enrolled. The volunteers included bush meat hunters, their family members, local health care workers and the general population of the village. Volunteers under 16 years of age, pregnant women, travellers or visitors, and people suffering from chronic disease were excluded from the original study. In total, 516 samples from the Macenta cohort were analysed. For the purpose of the analysis, 68 samples from Guéckédou city, collected in 2018 in the context of a longitudinal study of EVD survivors, were included in the Macenta population. Both Guéckédou city and Macenta prefecture are forested regions. All samples were stored at - 80°C in a temperature monitored unit within the PHE Porton Down UK laboratories. Consent forms stating the study aims and their rights as volunteers were signed prior to sample collection for each individual study.

### Blood sample collection and processing

A volume (5-20ml) of human peripheral blood was collected into non-anticoagulant BD Vacutainer® blood collection tubes from participants. Blood clotted within two hours and transported to the laboratory in a cool box at 4°C. Samples were centrifuged at 2,000 x g for 10 minutes at room temperature and serum aliquoted into 2 ml microtube RT-PCR tubes (Starsted, REF 72.694.406) in an air purifying class II microbiological safety cabinet (Envair Ltd, England). Aliquots of sera were shipped on dry ice at -20°C and processed at PHE Porton Down laboratories (England).

### Samples analysis by LASV-specific ELISA

To measure anti-LASV IgG specific antibodies in sera, the patented BLACKBOX® LASV IgG kit from Bernhard Nocht Institute for Tropical Medicine (BNITM) in Germany (ELG.004) used for qualitative serological detection of acute or past LASV infection *(21) (22)*. The kit was used as per the manufacturer’s instructions. Each sample was run as single replicate. All reagents were provided by BNITM. Positive and negative controls were provided with the kit. Blank and background wells were added to monitor non-specific bindings of proteins to reagent contaminants or to the ELISA plate surface. Briefly, initial plate washing was performed using pre-diluted wash buffer. Then, 25uL of diluted biotinylated recombinant LASV antigen (nucleoprotein) were transferred onto the ELISA plate and 25uL of diluted control or patient sample added prior to plate incubation for 24h at 4°C in a humid chamber. After a wash step, 50uL of diluted streptavidin/Horseradish Peroxidase (HRP) were added and plate incubated for 1h at 4°C. Another wash step was performed, 100uL/well of 3,3’,5,5’-Tetramethylbenzidine (TMB) peroxidase substrate were added and plate incubated for 10min at room temperature in the dark. Finally, 100uL/well of Stop solution (1M H2SO4) were added and optical density (OD) readings (450-620nm) obtained by using SpectraMax plate reader (software SoftMaxPro v9). As recommended in the BLACKBOX® LASV IgG ELISA manual, results were interpreted as valid if the following criteria were met: OD450-OD620 of negative control < 0.10 and OD450-OD620 of positive control > 1.00. Average absolute OD450-OD620 absorbance value for negative control samples (*OD*_*Neg,AV*_) was used to calculate the assay cut off *OD*_*CO*_ according to the following formula:

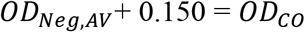

Index Values (IV) for the tested samples were calculated as follows:

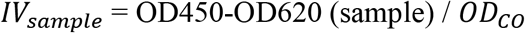

Results classification for index values was carried out as follows:

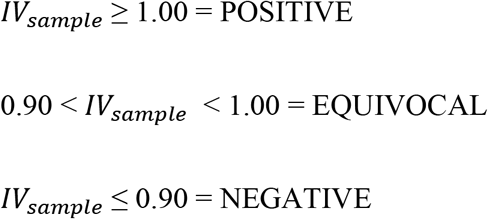

### Environmental and spatial covariates

To assess the impact of land cover and other environmental covariates on LASV exposure, we assembled a dataset of remote-sensing derived land cover maps from 2017 at 100m resolution *(23)*. We reclassified land cover into the following categories: closed forest (canopy cover >70%), open forest (canopy cover 15-70%), shrubs, herbaceous vegetation (plants without persistent stems and canopy cover <10%) and urban/built up areas. We additionally extracted data on the percentage canopy cover for all sites. Population density was obtained from village level population estimates of all sampled areas.

As sampled villages were only geolocated to centroids, we extracted environmental data within buffer radii of 500m, 1km, 2km, 5km and 10km from the village centroid to represent the possible distributions of the village and lands used *(24)*. For all buffer radii, we extracted the proportions of different land cover types and the mean canopy cover. To evaluate the importance of landscape configuration, we additionally extracted metrics on fragmentation, including perimeter area ratio, shape index (patch perimeter divided by the minimum possible patch perimeter) and fractal dimension index (an index of shape complexity) *(25)*. In ecology, fragmentation is defined as the breaking up of habitat and it is often used as a measurable index, usually related to human activity, which produces loss or reduction in habitat size or increased isolation of habitats.

### Statistical analyses

To characterise transmission intensity in both sites, we first graphed both continuous antibody responses and age-specific estimates of seropositivity. Reversible catalytic models were fit to age sero-prevalence data using maximum likelihood methods *(15)*. We used profile likelihood plots to explore possible historical changes in transmission intensity, with final models selected using likelihood ratio tests *(26)*. Final models were used to estimate seroconversion rates (SCR).

We additionally assessed whether fine-scale patterns of serological exposure were associated with landscape characteristics. As environmental variables can be associated with risk at different spatial scales, we used a data-driven approach to identify environmental risk factors *(27)*. First, mixed-effects logistic regression models were run for each environmental variable at each spatial scale (126 in total) to identify factors associated with LASV seropositivity. Models were run separately for both sites, with the primary outcome as the binary classification for individual level seropositivity. Models were fit with village as a random effect to account for observations within villages lacking independence. We first assessed associations with key demographic factors, including gender, continuous age and age category. For the Macenta/Guéckédou (forested region) model, only environmental predictors were included as demographic factors (age, gender) were not associated with exposure risks. For the coastal site, age category was included in all models in addition to environmental predictors. When assessing for inclusion in the multivariate analysis, univariate results were considered significant at p < 0.2 to avoid exclusion of variables that alone lack significance but contribute in presence of other variables. Highly correlated variables (Pearson’s correlation coefficient > 0.75) were removed. Where environmental covariates appeared significantly associated with the outcome of interest across multiple buffer radii, inclusion in the final model was based on Akaike’s Information Criteria (AIC). Final multivariate models were fit in a forward stepwise manner using AIC, with variables considered significant at p < 0.05.

## Results

Within this study, we assessed LASV IgG seropositivity in 1286 individuals using serum samples collected from the period of 2016 to 2018 in Guinea. In the Coastal site, 702 samples were analysed from 7 prefectures, encompassing 24 subprefectures and 29 villages/neighbourhoods (**Figure 1A, Table 1, Appendix Table 1**). In the Forested site, 516 samples were analysed from Macenta prefecture and 68 from Guéckédou city. The Macenta samples originated from 7 subprefectures encompassing 44 villages (**Figure 1A, Table 1, Appendix Table 2**).

**Table 1:** Study participants of coastal and forested regions.

| Coastal site |  | Forested Region |  |
| --- | --- | --- | --- |
| N | **702** | N | **584** |
|  |  | Macenta | 516 |
|  |  | Guéckédou | 68 |
| Female | 243 (34.6%) | Female | 262 (44.9%) |
| Male | 459 (65.4%) | Male | 322 (51.1%) |
| Age range (years) | 6-99 | Age range (years) | 17-90 |
| Mean age (years) | 32.3 | Mean age (years) | 38.8 |
| Sampling dates | 2016 | Sampling dates | 2017-2018 |

We observed that exposure to LASV was highly heterogenous both between (**Figure 1A**) and within (**Figure 1B**) each study site. The LASV IgG seropositivity was 11.9% (95% CI 9.7-14.5) in the Coastal site (**Figure 1B, Appendix Table 1**), while it was 59.6% (95% CI 55.5-63.5) in the Forested site (**Figure 1B, Appendix Table 2**).

Demographic characteristics of exposed individuals varied by study site. In the Coastal site, seropositivity in females was 9.0% (95% CI 5.8-13.1) and in males was 13.5% (95% CI 11.0-17.0). For the Forested site, seropositivity in females was 61.8% (95% CI 56.0-67.7) and in males was 58% (95% CI 52.5-63.3). Gender was not significantly associated with seropositivity in either site.

Age patterns of seropositivity differed markedly between sites. In the Coastal site, exposure was strongly associated with age, with seropositivity increasing with age (**Figure 2A**). However, no correlation was found between the quantitative antibody responses expressed by Index Value and age among seropositive individuals (**Appendix Figure 1**). Within this population, reversible catalytic models estimated the force of infection (Seroconversion rate = SCR) as 0.0041 (95% CI: 0.0033 – 0.0050). No changes in historical transmission intensity were detected (*p* = 0.328). In contrast, within the Forested site, no clear age patterns were identified and neither continuous nor categorical age were associated with individual seropositivity (**Figure 2B**). Although age ranges for included individuals were different between both sites (**Table 1**), substantial differences in exposure risks were detected in similar age groups between sites. For example, individuals aged 17-20 had an estimated seropositivity of 5.4% (95% CI: 1.5-13.3%) in the Coastal site compared to 50.0% in the Forested site (95% CI: 37.2 – 62.8%). The high proportions of seropositivity in the youngest age groups and lack of association with age precluded fitting reversible catalytic models for the Forested region.

**Figure 2.**
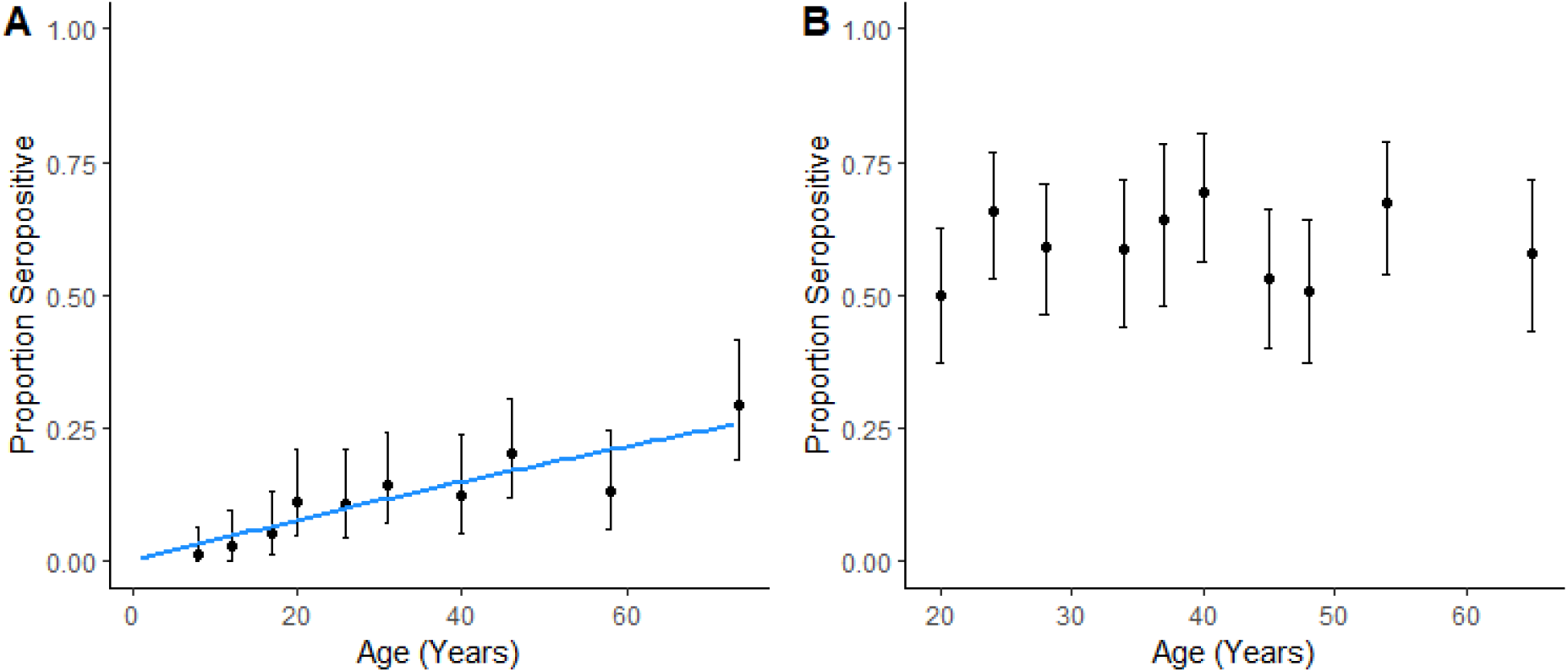
Proportion seropositive with 95% Confidence intervals by age decile for Coastal site (**A**) and Forested site (**B**), blue line is modelled SCR curve.

Despite clear differences in transmission between sites, strong associations between exposure and environmental factors were identified for both sites. The land cover classes for both sites are shown in **Figure 3A**. As characterised by perimeter area ratios, fragmentation was strongly positively associated with exposure risk for both sites (**Figure 3B and C, Appendix Tables 3 and 4**). In the Coastal site, the proportion of open forests and fragmentation of built up areas increased odds of exposure. Other land cover factors, such as closed forests and vegetation fragmentation, were not significantly associated with exposure risks but improved overall model fit (**Figure 3B, Appendix Table 3**). Within the Forested site, increased fragmentation of both open and closed forests was positively associated with risk, while fragmented vegetation was protective (**Figure 3C, Appendix Table 4**). Across both sites, significant variables were identified at different spatial scales, consistent with other studies of environmental risk factors for zoonotic diseases *(28)*.

**Figure 3:**
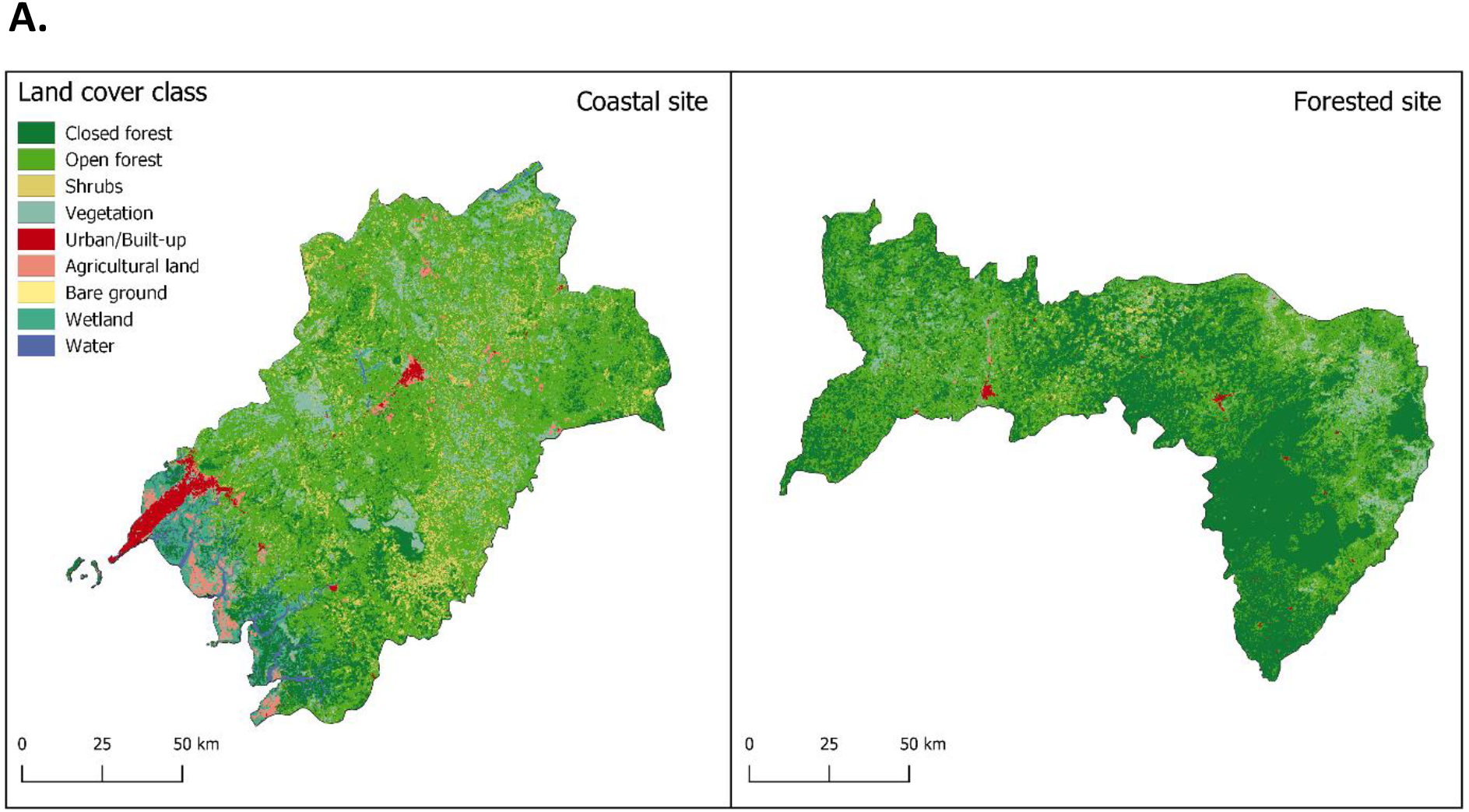
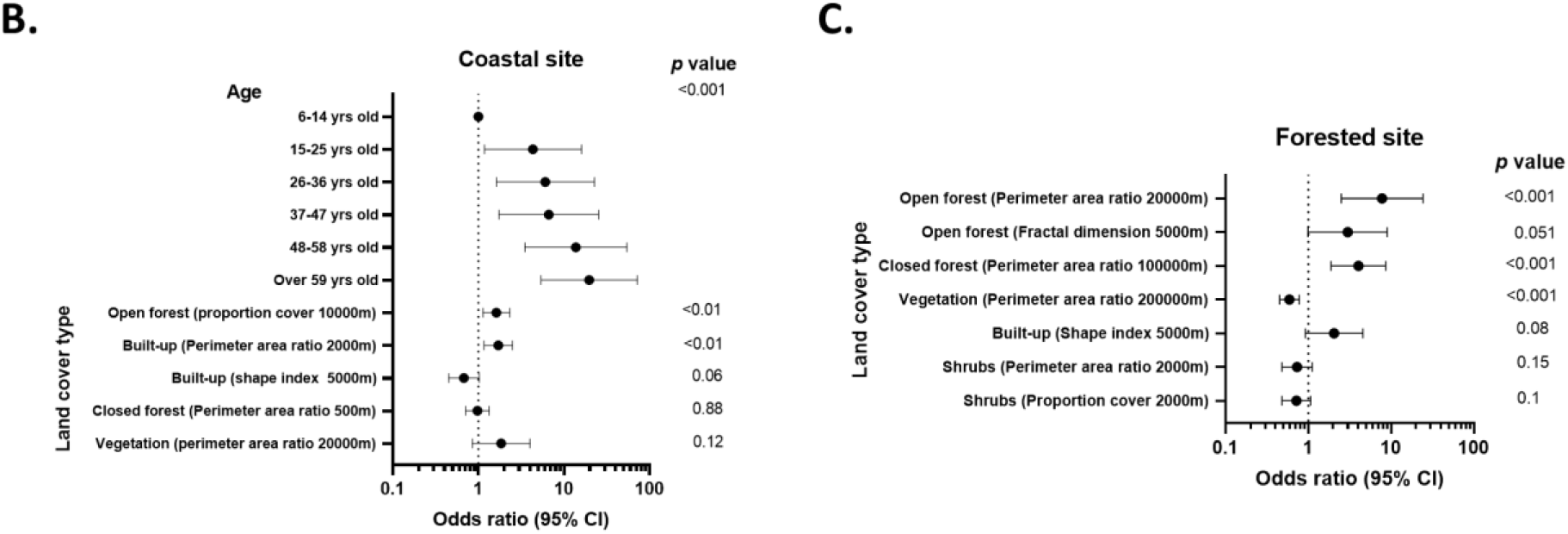
**A)** Land cover classes for the Coastal and Forested sites. **B and C**) Associations between exposure risk and environmental factors in the Coastal (**B**) and Forested sites (**C**). OR: Odd ratios. CI: Confidence intervals.

## Discussion

Despite the growing interest in Lassa fever, there is limited up-to-date information regarding the seropositivity of LASV in West Africa. To our knowledge, this study is the first one to estimate transmission intensity from serological data in Guinea and analyse the environmental risk factors. We found that LASV NP-specific IgG seropositivity differed between our study sites: Coastal and Forested regions. In 1993, Lukashevich and colleagues had also found a high LASV NP-specific IgG positivity among inhabitants of tropical secondary forest (areas near Guéckédou, Yomou, and Lola) and guinea savannah (Faranah and Kindia areas) (25-55%) and a lower positivity among inhabitants of mountainous (Pita, Labe, and Mali) and coastal areas (Boffa, Boké) (4-7%) *(18)*. In our study, we found that gender was not significantly associated with seropositivity in either site, which is in line with other studies conducted both in Guinea and Sierra Leone *(18) (12)*. Interestingly, our findings showed a significant increase in LASV IgG seropositivity with age in the Coastal site but not in the Forested site. Although in the above-mentioned study, Lukashevich and colleagues did not detect any correlation between antibody prevalence and age in any studied regions *(18)*, differences in age patterns of seropositivity between regions were observed in other studies in West Africa. A study conducted in Forested Guinea in the year 2000, found individuals under 10 and between 20 and 29 years of age to have the highest LASV IgG prevalence *(7)*. A study conducted in Sierra Leone, found antibody prevalence to increase with age, reaching a peak at 20-50 years of age and decreasing thereafter *(12)*. A similar trend was reported in another pilot study conducted in 2015 in Nigeria *(29)*. Age, often associated with occupation or daily activities, may play a role on exposure but our results in the forested region and others studies suggest that in some regions, other factors such as environmental features may have a bigger impact on exposure risk. It is also possible that more localised exposure within the household plays a role for viral transmission. Finally, we found an association between landscape patterns and exposure risk for both study sites. Associations between landscape fragmentation and exposure risk to zoonotic diseases have been observed for other zoonotic diseases such as malaria *(28) (30)* and Ebola virus *(31)*. In our study, the fragmentation of forests and their proportion appear to have an impact on exposure risk but their exact role in the complex dynamics of LASV circulation remains to be determined.

The murine population is another variable involved in the distribution of LASV. In one study looking at LASV-specific IgG prevalence in the reservoir host *M. natalensis* in Guinea, the authors found that 0% of the M. *natalensis* were infected with Lassa virus in the low seroprevalence area (the coastal region) while 5.4%-32.1 % of sampled mice were infected with the virus in the high seroprevalence region (forested guinea) *(10)*. Another study sampled a total of 1616 small rodents (956 *Mastomys* sp.) captured in 444 households in the savanna, mixed savanna-forest and forest region of Guinea were studied for LASV-specific antibody and antigen detection. The authors found that the proportion of LASV infected rodents per region ranged from 0% - 9%. It was higher in the savannah and forested areas. Interestingly, they observed that the proportion of infected animals in each village was highly variable amongst neighboring villages, identifying ‘hot spots’ of LASV infected mice clustering within few homes, thus adding a further element that could drive the highly heterogeneous distribution of LASV *(32)*. Even though the distribution of *M. natalensis* is an important factor, its presence may not be sufficient to explain the IgG seropositivity in humans *(33) (10)*, as additional rodent reservoirs have been identified revealing the potential for a wider ecological niche for LASV *(5)*.

Finally, some limitations of our study can be highlighted. The type of ELISA used to assess LASV seropositivity may impact on the results. Even though the BLACKBOX IgG ELISA was shown to be sensitive and very specific (90% and 99%) when compared to the immunofluorescence assay *(21)*, it was not tested against other arenaviruses due to lack of suitable serum panels. Consequently, we cannot fully exclude the potential circulation of related, or perhaps yet undescribed arenaviruses, in specific regions *(34)* and a partial cross-reactivity of IgG antibodies against other arenaviruses. In addition, as the samples were collected as part of a vaccine evaluation and a sero-epidemiology study on Ebola virus, the sampling procedure was not standardised between the studies. It would have been useful to have additional information on individual’s LASV exposure risks i.e. occupation, relevant medical history, hunting/foraging habits, location of previous residence, methods for food storage and rodents trapping around the house. It would be useful to know the number of people in the household, the quality of the building, as well as presence of rodent burrows in or around the house. Finally, this study measured only IgG antibodies in sera collected at one timepoint. As such, our data does not allow us to capture the potential effect of changes in seasonal rodent activity or vegetation patterns surrounding the villages. Also, since people tend to move frequently across wide regions, either to find work during harvest season or to hunt *(35)* and some may have moved away from Ebola virus disease hotspots during the 2013-2016 outbreak, they may have been exposed to LASV in a different location from the one in which they resided at the time of sampling.

Similar to other endemic tropical diseases, LASV dynamics result from complex interactions between several human and environmental factors. Our study compared the LASV-specific IgG seropositivity in Coastal and Forested regions in Guinea and for the first time described in detail association between exposure to LASV and landscape patterns. Enhanced knowledge of regions with an a potentially increased exposure risk to LASV can help to improve the epidemiological surveillance, devise one health interventions and highlight potential trial sites to assess new LASV vaccines and therapeutics.

## Data Availability

All data produced in the present study are available upon reasonable request to the authors

## Acknowledgments

This work was funded by the US Food & Drug Administration (HHSF223201510104C) and the CGIAR Research Program on Agriculture for Nutrition and Health (A4NH). SG was supported by grants GU883/5-1 and GU883/5-2 from the German Research Foundation (DFG). KMF is supported by a Sir Henry Dale Fellowship jointly funded by the Wellcome Trust and the Royal Society (Grant no. 221963/Z/20/Z).

## Notes

### Competing Interest Statement

The authors have declared no competing interest.

### Funding Statement

This study was funded by the US Food & Drug Administration (HHSF223201510104C) and the CGIAR Research Program on Agriculture for Nutrition and Health (A4NH). SG was supported by grants GU883/5-1 and GU883/5-2 from the German Research Foundation (DFG). KMF is supported by a Sir Henry Dale Fellowship jointly funded by the Wellcome Trust and the Royal Society (Grant no. 221963/Z/20/Z).

### Author Declarations

The London School of Hygiene and Tropical Medicine Ethics committee (Ethics Ref: 17429) gave ethical approval for this work. The Comite national d ethique pour la recherche en sante (CNERS) of Republic of Guinea gave ethical approval for this work on the 16th of March 2017 (012/CNERS/17).

## References

1. Hensley, L.E.; Smith, M.A.; Geisbert, J.B.; Fritz, E.A.; Daddario-DiCaprio, K.M.; Larsen, T.; et al. Pathogenesis of Lassa fever in cynomolgus macaques. Virol J 2011, 8, 205.

2. Richmond, J.K.; Baglole, D.J. Lassa fever: epidemiology, clinical features, and social consequences. BMJ 2003, 327, 1271–1275.

3. McCormick, J.B.; Walker, D.H.; King, I.J.; Webb, P.A.; Elliott, L.H.; Whitfield, S.G.; et al. Lassa virus hepatitis: a study of fatal Lassa fever in humans. Am J Trop Med Hyg 1986, 35, 401–407.

4. Monath, T.P.; Newhouse, V.F.; Kemp, G.E.; Setzer, H.W.; Cacciapuoti, A. Lassa virus isolation from Mastomys natalensis rodents during an epidemic in Sierra Leone. Science 1974, 185, 263–265.

5. Olayemi, A.; Oyeyiola, A.; Obadare, A.; Igbokwe, J.; Adesina, A.S.; Onwe, F.; et al. Widespread arenavirus occurrence and seroprevalence in small mammals, Nigeria. Parasit Vectors 2018, 11, 416.

6. Fisher-Hoch, S.P.; Tomori, O.; Nasidi, A.; Perez-Oronoz, G.I.; Fakile, Y.; Hutwagner, L.; et al. Review of cases of nosocomial Lassa fever in Nigeria: the high price of poor medical practice. BMJ 1995, 311, 857–859.

7. Kerneis, S.; Koivogui, L.; Magassouba, N.; Koulemou, K.; Lewis, R.; Aplogan, A.; et al. Prevalence and risk factors of Lassa seropositivity in inhabitants of the forest region of Guinea: a cross-sectional study. PLoS Negl Trop Dis 2009, 3, e548.

8. Stephenson, E.H.; Larson, E.W.; Dominik, J.W. Effect of environmental factors on aerosol-induced Lassa virus infection. J Med Virol 1984, 14, 295–303.

9. Ajayi, N.A.; Nwigwe, C.G.; Azuogu, B.N.; Onyire, B.N.; Nwonwu, E.U.; Ogbonnaya, L.U.; et al. Containing a Lassa fever epidemic in a resource-limited setting: outbreak description and lessons learned from Abakaliki, Nigeria (January-March 2012). Int J Infect Dis 2013, 17, e1011–1016.

10. Lecompte, E.; Fichet-Calvet, E.; Daffis, S.; Koulemou, K.; Sylla, O.; Kourouma, F.; et al. Mastomys natalensis and Lassa fever, West Africa. Emerg Infect Dis 2006, 12, 1971–1974.

11. Ogbu, O.; Ajuluchukwu, E.; Uneke, C.J. Lassa fever in West African sub-region: an overview. J Vector Borne Dis 2007, 44, 1–11.

12. McCormick, J.B.; Webb, P.A.; Krebs, J.W.; Johnson, K.M.; Smith, E.S. A prospective study of the epidemiology and ecology of Lassa fever. J Infect Dis 1987, 155, 437–444.

13. Gibb, R.; Moses, L.M.; Redding, D.W.; Jones, K.E. Understanding the cryptic nature of Lassa fever in West Africa. Pathog Glob Health 2017, 111, 276–288.

14. Bond, N.; Schieffelin, J.S.; Moses, L.M.; Bennett, A.J.; Bausch, D.G. A historical look at the first reported cases of Lassa fever: IgG antibodies 40 years after acute infection. Am J Trop Med Hyg 2013, 88, 241–244.

15. Drakeley, C.J.; Corran, P.H.; Coleman, P.G.; Tongren, J.E.; McDonald, S.L.; Carneiro, I.; et al. Estimating medium-and long-term trends in malaria transmission by using serological markers of malaria exposure. Proc Natl Acad Sci U S A 2005, 102, 5108–5113.

16. Greenhouse, B.; Daily, J.; Guinovart, C.; Goncalves, B.; Beeson, J.; Bell, D.; et al. Priority use cases for antibody-detecting assays of recent malaria exposure as tools to achieve and sustain malaria elimination. Gates Open Res 2019, 3, 131.

17. Arnold, B.F.; Scobie, H.M.; Priest, J.W.; Lammie, P.J. Integrated Serologic Surveillance of Population Immunity and Disease Transmission. Emerg Infect Dis 2018, 24, 1188–1194.

18. Lukashevich, I.S.; Clegg, J.C.; Sidibe, K. Lassa virus activity in Guinea: distribution of human antiviral antibody defined using enzyme-linked immunosorbent assay with recombinant antigen. J Med Virol 1993, 40, 210–217.

19. Bausch, D.G.; Demby, A.H.; Coulibaly, M.; Kanu, J.; Goba, A.; Bah, A.; et al. Lassa fever in Guinea: I. Epidemiology of human disease and clinical observations. Vector Borne Zoonotic Dis 2001, 1, 269–281.

20. Thom, R.; Tipton, T.; Strecker, T.; Hall, Y.; Akoi Bore, J.; Maes, P.; et al. Longitudinal antibody and T cell responses in Ebola virus disease survivors and contacts: an observational cohort study. Lancet Infect Dis 2021, 21, 507–516.

21. Gabriel, M.; Adomeh, D.I.; Ehimuan, J.; Oyakhilome, J.; Omomoh, E.O.; Ighodalo, Y.; et al. Development and evaluation of antibody-capture immunoassays for detection of Lassa virus nucleoprotein-specific immunoglobulin M and G. PLoS Negl Trop Dis 2018, 12, e0006361.

22. Schmitz, H.; Gabriel, M.; Emmerich, P. Specific detection of antibodies to different flaviviruses using a new immune complex ELISA. Med Microbiol Immunol 2011, 200, 233–239.

23. Buchhorn, M.; Smets, B.; Bertels, L.; De Roo, B.; Lesiv, M.; Tsendbazar, N.-E.; et al. Copernicus Global Land Service: Land Cover 100m: collection 3: epoch 2019: Globe. Zenodo 2020.

24. Brock, P.M.; Fornace, K.M.; Grigg, M.J.; Anstey, N.M.; William, T.; Cox, J.; et al. Predictive analysis across spatial scales links zoonotic malaria to deforestation. Proc Biol Sci 2019, 286, 20182351.

25. McGarigal, K.; Cushman, S.; Catherine Neel, M.; E., E. Spatial Pattern Analysis Program for Categorical Maps. Available online: http://www.umass.edu/landeco/research/fragstats/fragstats.html

26. Stewart, L.; Gosling, R.; Griffin, J.; Gesase, S.; Campo, J.; Hashim, R.; et al. Rapid assessment of malaria transmission using age-specific sero-conversion rates. PLoS One 2009, 4, e6083.

27. Fornace, K.M.; Brock, P.M.; Abidin, T.R.; Grignard, L.; Herman, L.S.; Chua, T.H.; et al. Environmental risk factors and exposure to the zoonotic malaria parasite Plasmodium knowlesi across northern Sabah, Malaysia: a population-based cross-sectional survey. Lancet Planet Health 2019, 3, e179–e186.

28. Stefani, A.; Dusfour, I.; Correa, A.P.; Cruz, M.C.; Dessay, N.; Galardo, A.K.; et al. Land cover, land use and malaria in the Amazon: a systematic literature review of studies using remotely sensed data. Malar J 2013, 12, 192.

29. Tobin, E.; Asogun, D.; Akpede, N.; Adomeh, D.; Odia, I.; Gunther, S. Lassa fever in Nigeria: Insights into seroprevalence and risk factors in rural Edo State: A pilot study. J Med Trop 2015, 17, 51–55.

30. Fornace, K.M.; Abidin, T.R.; Alexander, N.; Brock, P.; Grigg, M.J.; Murphy, A.; et al. Association between Landscape Factors and Spatial Patterns of Plasmodium knowlesi Infections in Sabah, Malaysia. Emerg Infect Dis 2016, 22, 201–208.

31. Olivero, J.; Fa, J.E.; Real, R.; Marquez, A.L.; Farfan, M.A.; Vargas, J.M.; et al. Recent loss of closed forests is associated with Ebola virus disease outbreaks. Sci Rep 2017, 7, 14291.

32. Demby, A.H.; Inapogui, A.; Kargbo, K.; Koninga, J.; Kourouma, K.; Kanu, J.; et al. Lassa fever in Guinea: II. Distribution and prevalence of Lassa virus infection in small mammals. Vector Borne Zoonotic Dis 2001, 1, 283–297.

33. Marien, J.; Kourouma, F.; Magassouba, N.; Leirs, H.; Fichet-Calvet, E. Movement Patterns of Small Rodents in Lassa Fever-Endemic Villages in Guinea. Ecohealth 2018, 15, 348–359.

34. Coulibaly-N’Golo, D.; Allali, B.; Kouassi, S.K.; Fichet-Calvet, E.; Becker-Ziaja, B.; Rieger, T.; et al. Novel arenavirus sequences in Hylomyscus sp. and Mus (Nannomys) setulosus from Cote d’Ivoire: implications for evolution of arenaviruses in Africa. PLoS One 2011, 6, e20893.

35. Mari Saez, A.; Cherif Haidara, M.; Camara, A.; Kourouma, F.; Sage, M.; Magassouba, N.; et al. Rodent control to fight Lassa fever: Evaluation and lessons learned from a 4-year study in Upper Guinea. PLoS Negl Trop Dis 2018, 12, e0006829.

