## Supplementary data for "Influence of landscape patterns on the exposure of LASV across diverse regions within the Republic of Guinea"

*Contributed equally to first authorship

^Corresponding author

**Appendix Table 1:** LASV IgG seropositivity in the Coastal site.

| Location | No. Samples (N) | No. Positives | Seropositivity (95% CI) |
|---|---|---|---|
| Coastal | 702 | 84 | 11.9 (9.7-14.5) |

| Prefectures | No. Samples (N) | No. Positives | Seropositivity (95% CI) |
|---|---|---|---|
| Boke | 40 | 5 | 12.5 (2.3 - 22.8) |
| Conakry | 100 | 2 | 2.0 (0 - 4.7) |
| Coyah | 27 | 0 | 0 |
| Dubreka | 84 | 2 | 2.4 (0 – 5.6) |
| Forecariah | 406 | 67 | 16.5 (12.9 – 20.1) |
| Fria | 10 | 1 | 10.0 (0 – 28.6) |
| Kindia | 35 | 7 | 20.0 (6.8 – 33.3) |

**Appendix Table 2:** LASV IgG seropositivity in the Forested site (Macenta/Guéckédou).

| Location | No. Samples (N) | No. Positives | Seropositivity (95% CI) |
|---|---|---|---|
| Forested | 584 | 348 | 59.6 (55.5-63.5) |

| Subprefectures | No. Samples (N) | No. Positives | Seropositivity (95% CI) |
|---|---|---|---|
| Binikala | 48 | 27 | 56.25 (41.18-70.52) |
| Bofossou | 190 | 150 | 78.95 (72.46-84.51) |
| Coyamah | 42 | 21 | 50.00 (34.19-65.81) |
| Fassankoni | 76 | 66 | 86.84 (77.13-93-51) |
| Oremai | 76 | 16 | 21.05 (12.54-31.92) |
| Seredou | 71 | 12 | 16.90 (9.05-27.66) |

| | | | |
|---|---|---|---|
| Wattanka | 13 | 10 | 76.92 (46.19-94.96) |
| Gueckedou | 68 | 46 | 67.65 (55.21-78.49) |

**Appendix Figure 1:** Quantitative IgG responses among seropositive individuals in the Coastal site. Spearman r = 0.03522, *P* value=0.7293.

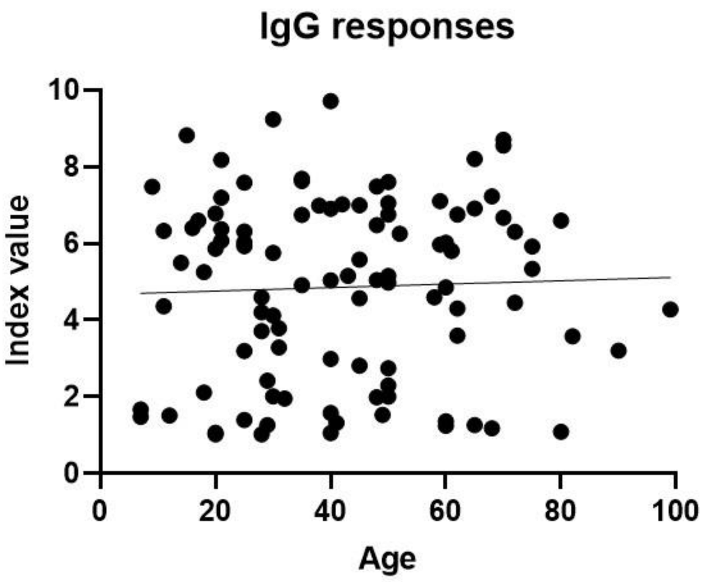


**Appendix Table 3:** Associations between exposure risk and environmental factors in the Coastal site. OR: Odd ratios.

| Variable | OR | 95% CI | p value |
|---|---|---|---|
| **Age category** | | | |
| **6-14** | **Baseline** | | |
| **15-25** | **4.332991** | **(1.18-15.96)** | **< 0.05** |
| **26-36** | **6.0496545** | **(1.62-22.6)** | **< 0.01** |

| | | | | | |
|---|---|---|---|---|---|
| 37-47 | | | 6.6435946 | (1.74-25.4) | < 0.01 |
| 48-58 | | | 13.764193 | (3.49-54.21) | < 0.001 |
| Over 59 | | | 19.695971 | (5.39-72) | < 0.001 |
| **Land cover type** | **Metric** | **Scale** | | | |
| Built-up | Perimeter:area ratio | 2000m | 1.7058627 | (1.16-2.5) | < 0.01 |
| Closed forest | Perimeter:area ratio | 500m | 0.975557 | (0.71-1.34) | 0.88 |
| Open forest | Proportion land cover type | 10000m | 1.6235948 | (1.13-2.32) | < 0.01 |
| Built-up | Shape index | 5000m | 0.6750841 | (0.45-1.02) | 0.06 |
| Vegetation | Perimeter:area ratio | 20000m | 1.8428392 | (0.85-4) | 0.12 |

**Appendix Table 4:** Associations between exposure risk and environmental factors in the Forested site. OR: Odd ratios.

| Land cover type | Metric | Scale | OR | 95% CI | p value |
|---|---|---|---|---|---|
| Open forest | Fractal dimension | 5000m | 2.99 | (0.99-8.99) | 0.051 |
| Open forest | Perimeter:area ratio | 20000m | 7.79 | (2.5-24.27) | < 0.001 |
| Built-up area | Shape index | 5000m | 2.05 | (0.92-4.59) | 0.08 |
| Shrubs | Perimeter:area ratio | 2000m | 0.73 | (0.48-1.12) | 0.15 |

| | | | | | |
|---|---|---|---|---|---|
| **Closed forest** | **Perimeter:area ratio** | **100000m** | **4.03** | **(1.88-8.63)** | **< 0.001** |
| **Vegetation** | **Perimeter:area ratio** | **200000m** | **0.59** | **(0.45-0.78)** | **< 0.001** |
| **Shrubs** | **Proportion of land cover** | **2000m** | **0.72** | **(0.48-1.07)** | **0.1** |
